# Test-Retest Reliability of an Isometric Trunk Flexor and Extensor Test in Community-Dwelling Adults

**DOI:** 10.64898/2026.09.21.26361499

**Authors:** Clare Dal Bon, Xanthe Meehan, Chelsea Lehmann, Patrick Beaumont, Paul Licina

## Abstract

**Background:** Trunk flexor and extensor measurement tools are useful for measuring progress throughout the identification, treatment and management of spinal pathologies. Handheld dynamometry is popular amongst clinicians due to its convenience. However, poor standardisation of the testing position can lead to suboptimal intra-rater reliability, which consequently reduces the validity of the results. The VALD ForceFrame Strength Testing System is a portable and standardized hybrid strength-assessment system that quantifies strength and power outputs.

**Objective:** This study aims to assess the test-retest reliability of isometric trunk flexion and extension strength using the ForceFrame testing technology.

**Methods:** A total of 29 healthy participants undertook two testing sessions, each requiring them to perform maximal isometric trunk flexion and extension using the ForceFrame. Test re-test reliability was assessed using the Intraclass Correlation Coefficient (ICC), and the standard error of measure (SEM) was used to calculate the minimum detectable change.

**Results:** Excellent reliability was observed for both trunk flexion (*ICC* 0.92; 95% CI 0.83-0.96) and extension (*ICC* 0.93; 95% CI 0.86-0.97) tests, and the minimum detectable changes observed were 71.35N and 101.20N, respectively.

**Conclusions:** The results from this study suggest that the VALD force frame offers clinicians a reliable method of assessing change in trunk flexor and extensor strength.

## INTRODUCTION

Clinicians treating spinal pathology do not routinely test trunk strength, primarily because current objective measurement techniques have issues with objectivity, uniformity and quantification. However, trunk flexor (TF) and extensor (TE) measurement tools may be useful in identifying risk factors, planning treatment and measuring progress. Research has found that in older adults, training to improve overall trunk strength and standing balance is associated with marked improvements in the performance of activities of daily living (Hosseini et al., 2012; Granacher et al., 2013). Moreover, recreational and elite athletes presenting with chronic low back pain (CLBP) generate a reduced TE Force compared to their pain-free counterparts (Moreno Catalá et al., 2018). Predictive research has suggested that early identification and strengthening of the TEs may reduce the incidence and severity of CLBP (Cho et al., 2014). Inhibition of the TFs has been identified as a pre-existing risk factor for the development of low back pain (Cholewicki et al., 2005).

Literature commonly cites two measurement tools for isometric muscle testing. The first is the isokinetic dynamometer, which is regarded as the gold-standard for muscle testing (Martin et al., 2006; Stark et al., 2011; Chopp-Hurley et al., 2019; Muñoz-Bermejo et al., 2019; Parraca et al., 2022). Isokinetic dynamometers (typically seen in research institutions or hospital-based rehabilitation centres), measure the force applied by a muscle group moving at either a constant velocity, or in a static, isometric position. The machine provides a resistance that prevents the limb from accelerating more than the pre-programmed velocity. Although considered the gold-standard of measurement devices, their size and cost restrict their use in out-patient rehabilitation clinics. As an inexpensive, more convenient alternative, hand-held dynamometry (HHD) was introduced as a tool to evaluate muscle strength. Despite its popularity, handheld dynamometry is unsuitable for testing all measures of isometric strength (De Blaiser et al., 2018). Isometric strength testing via HHDs requires the examiner to apply a force greater than or equal to the maximum voluntary contraction (MVC) exerted by the participant (van der Ploeg and Oosterhuis, 1991). While such a method may be appropriate for movements that recruit smaller muscle groups – and where the examiner is able to overcome the force of the participant – problems arise when testing larger muscle groups. It has been purported that where muscle forces exceed >15kg HHDs are impractical unless additional patient stabilisation procedures are applied (Brinkmann, 1994). Furthermore, standardisation of the testing position and the examiner’s level of experience both contribute towards HHD test error (Andrews et al., 1996; Macfarlane et al., 2008; Schrama et al., 2014). Given these shortcomings, there is a role for a hybrid strength-assessment system that is portable, controls for clinician error and standardises the testing position.

A method designed to overcome this hurdle involves the external fixation of the HHD to a solid base of support. In doing so, researchers have demonstrated an increase in the participants’ strength output, whilst achieving good-to-excellent scores of test-retest reliability (Bohannon et al., 2012; Harding et al., 2017). One such piece of technology is the Force Frame Testing System (FFTS), developed by VALD Performance, Brisbane, Australia. Similar to the HHD, the FFTS quantifies strength and power outputs of specific muscle groups (Jones et al., 2021). The FFTS relies on a movable external frame with multiple fixation points to ease position standardisation, providing a strong external resistance and increasing the objectivity of results. Kadlec et al. (2021) used the FFTS to assess the test-retest reliability of isometric hip adductor and abductor strength, for which it demonstrated good to excellent test-retest reliability (*ICC*= 0.86-0.91 and *ICC*= 0.86-0.92, respectively). Separate research has shown that the FFTS has greater reliability than the HHD in measuring hip abductor strength (*ICC*= 0.81-0.84) (Malliaras et al., 2009). However, there is a paucity of data measuring the precision of isometric trunk flexion (TF_ISO_) and extension (TE_ISO_) strength using the FFTS.

The aim of this study is to assess the test-retest reliability of the TF_ISO_ and the TE_ISO_ using the FFTS technology in community-dwelling adults. The goal is to provide a direct objective measurement method that will deliver clinicians and patients with timely and accurate feedback on their TF_ISO_ and TE_ISO_ strength.

## METHODS

### Study design and equipment

A trunk flexion and extension test were performed to assess back extensor and trunk muscle strength. The VALD ForceFrame was initially designed for strength testing in high-performance athletes however, modification was necessary to ease the testing procedure for patients with prohibiting ailments. The first was securing the frame and dynamometer unit to the treatment bed (Figure 1). This modification provided a moveable base of support that was adjustable depending on individual participant requirements. As the device is used for evaluating muscle strength in other body regions, the ForceFrame modification made it possible to assess the large proportion of patients at the orthopaedic spine clinic who are unable to lie on the floor. Further modification was made to the head unit, where an additional sensor was incorporated into the front of the unit, enabling additional strength testing beyond the capabilities of the unaltered version. Of note, this modified ForceFrame is not yet commercially available. Prior to commencing testing a standardised warm-up protocol was administered. This included gentle back extensor, gluteal, hamstring and hip flexor stretches. Each testing session had an estimated duration of 15-minutes, inclusive of the warm-up protocol. Participants performed a single familiarisation trial for each test before completing one formal trial (Harding et al., 2017). The same researchers conducted both testing sessions.

**Figure 1.**
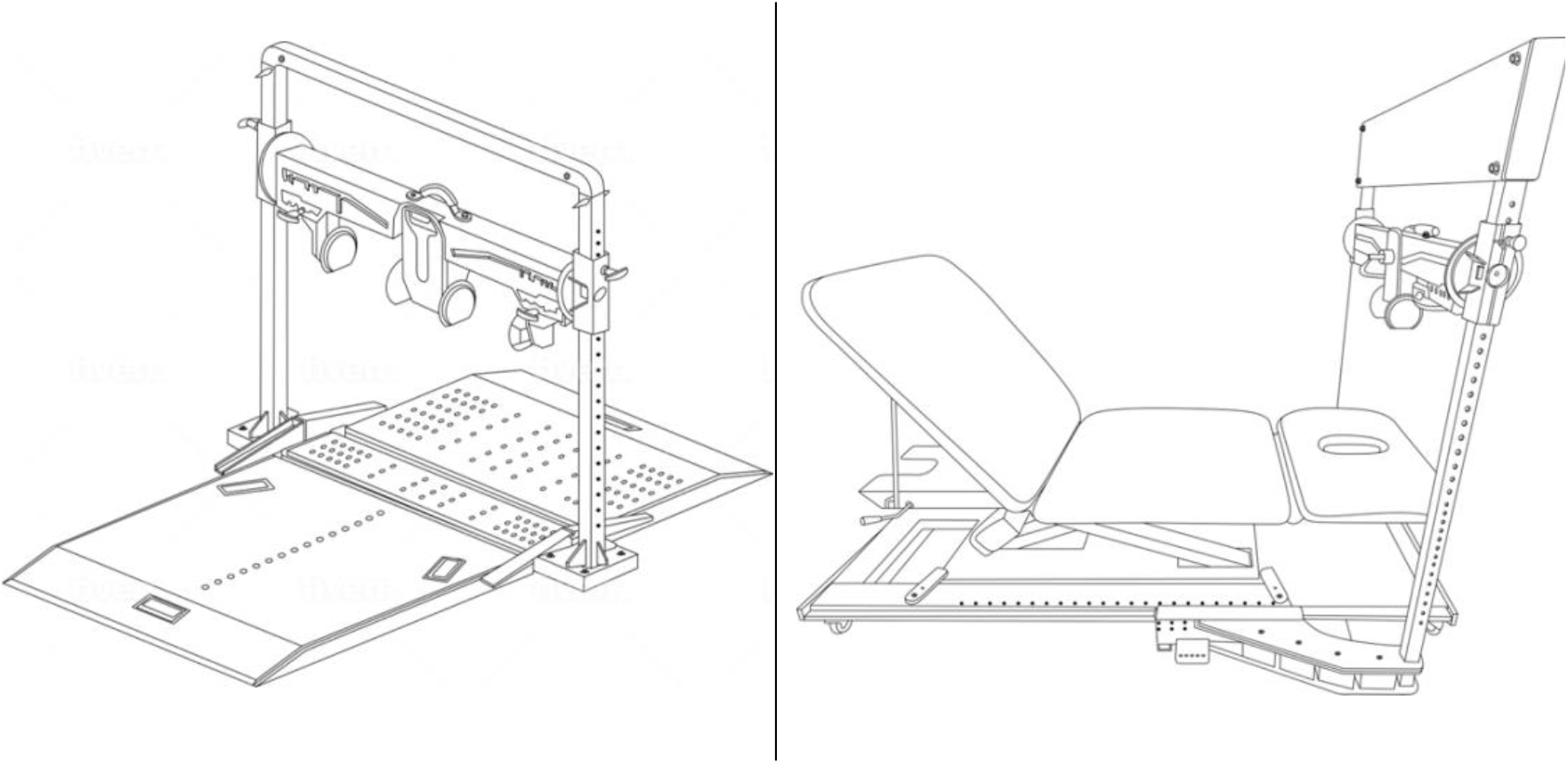
Unmodified VALD ForceFrame (left) compared with the custom-made ForceFrame (right) fitted to an examination bed.

### Isometric Trunk Flexion

The TF_ISO_ objectively measures the composite strength of the trunk flexors whilst limiting the action of synergist musculature. Prior to assuming the testing position, a small marker was placed on the sternum, 10cm below the suprasternal notch, providing the assessors with a consistent indicator for positioning the Force plate between trials (De Blaiser et al., 2018). Following this, the participant was instructed to sit on the examination bed with their legs straight, their hands resting on their thighs and their back resting against the raised bed backrest. The backrest was positioned at a -40° angle for both trials as this angle has demonstrated the greatest torque production in previous studies (Deering et al., 2017). The assessors then positioned the FFTS in line with the pre-placed chest marker. The participant was instructed to slide 20-cm forward, and another goniometer measurement was taken when the participant was re-positioned. The backrest remained parallel to the participant’s trunk position (Figure 2). The height of the ForceFrame crossbar and the horizontal displacement of the FFTS in relation to the treatment bed was recorded to ensure the standardisation of the testing position within participants. The assessor applied resistance to the participants’ feet and then instructed the participant to contact the force pad and apply minimal force. The instruction to provide minimal force was intended to reduce the force produced by the momentum of hinging forwards. The assessor then instructed the participant to make contact with the force pad where maximum force was achieved within the first two seconds of contact, before the force trace began to decrease and the participant was advised to relax. Verbal encouragement was provided by the examiners. The result of the MVC was recorded in Newtons (N) via the FFTS software.

**Figure 2.**
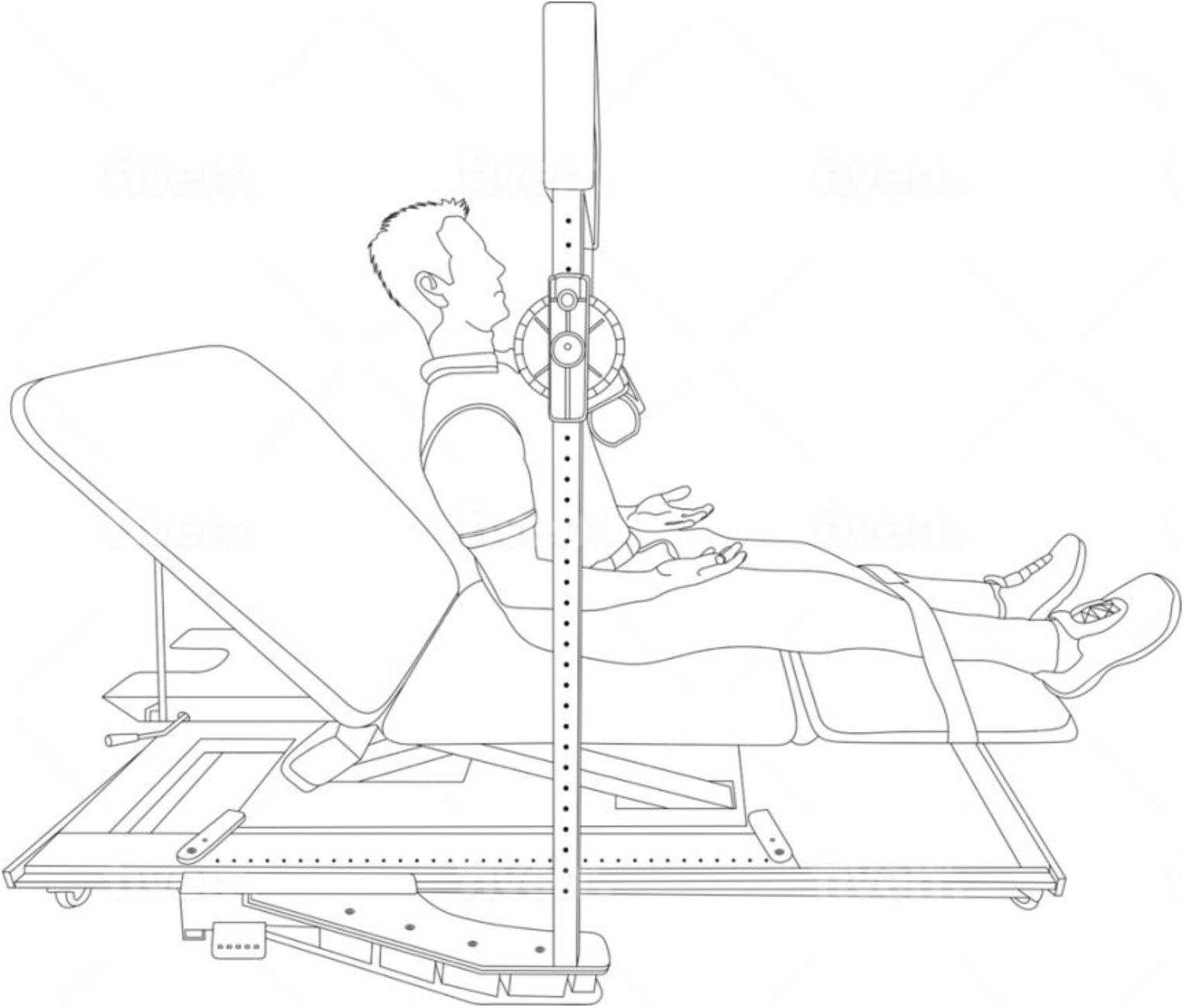
Representation of the trunk flexor testing position.

### Isometric Trunk Extension

The TE_ISO_ objectively measures the composite strength of the trunk extensors whilst limiting the action of synergist musculature. Prior to assuming the testing position, a small marker was placed 10cm below the spinous process of C7, at approximately the level of T4, providing the assessors with a consistent indicator for positioning the force plate between trials (De Blaiser et al., 2018). The participant was instructed to lie prone on the examination bed, with their hands resting on the back of their head. The FFTS was lowered into position, in line with the pre-placed marker (Figure 3). The height of the ForceFrame crossbar and the horizontal displacement of the FFTS in relation to the treatment bed was recorded to ensure the standardisation of the testing position within participants. The assessor applied resistance to the participants’ feet and then instructed the participant to apply maximal force to the force pad for five seconds by pushing up against the pad and lifting their head and elbows off the bed. Verbal encouragement was provided by the examiners. The result of the MVC was recorded in Newtons (N) via the FFTS software.

**Figure 3.**
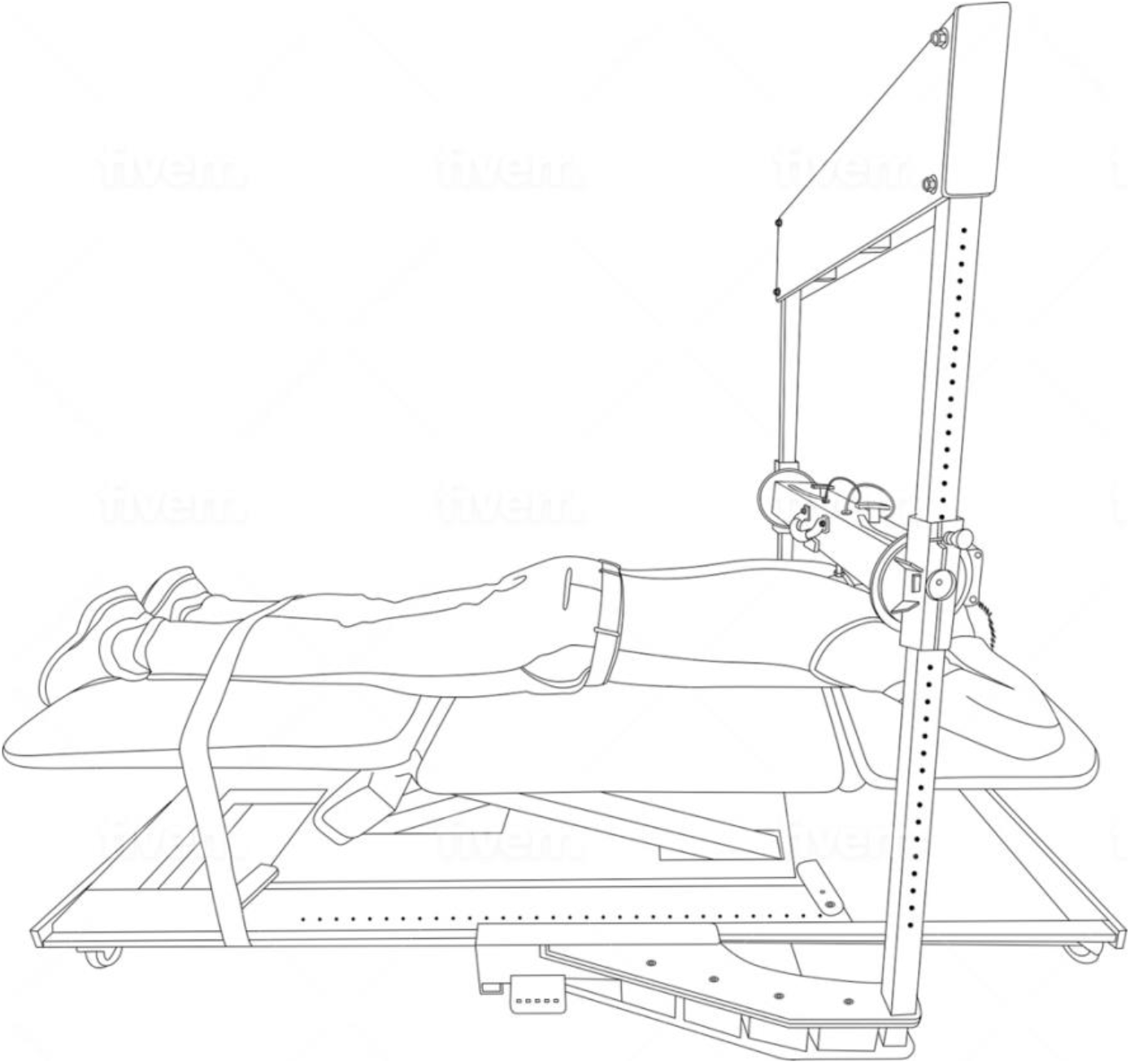
Representation of the trunk extensor testing position.

### Participants

Prior to recruitment, participants were screened using a musculoskeletal questionnaire. Upon completion of the questionnaire, participants were excluded from the study if they had an active musculoskeletal condition which could be exacerbated by the test or could influence the results of the trials. Participants were encouraged to continue their regular physical activity but asked to avoid trunk flexion and extension exercises between testing sessions, which were scheduled 48-hours to two weeks apart. Each subject was required to provide written consent prior to the commencement of testing (Brisbane Private Hospital Low Risk Ethics Review Committee – reference #LREC21BPH2).

### Statistical Analysis

The force (in Newtons) was collected from one maximal effort across two trials. The test-retest reliability was assessed using the method proposed by Shrout and Fleiss (1979). This method calculates the test-retest reliability using an Intraclass Correlation Coefficient (ICC) with a 95% confidence interval based on a two-way mixed-effects, single-rater, absolute agreement model (Koo and Li, 2016). Intraclass Correlation Coefficients of <0.5 were interpreted as poor, 0.5-0.75 moderate, 0.75-0.9 as good and >0.9 as excellent (Koo and Li, 2016). The ICC coefficients were calculated, reported, and interpreted for TF_ISO_ and the TE_ISO_. The trial-to-trial noise was calculated using the standard error of measurement (SEM) as: SEM= SD_b_ x

√(1-ICC) where SD_b_= standard deviation at baseline. The SEM% was calculated using the formula: (SEM/ mean) x 100 (Ransom et al., 2020). The SEM was then used to determine the minimum detectable change (MDC) at the 95% confidence interval considered as a true change in performance and was calculated using the formula: MDC_95_= 1.96 x √2 x SEM where 1.96 is the *z*-score for the 95% confidence interval. All analyses were conducted using SPSS Version 26 (Armonk, NY: IBM Corp.).

## RESULTS

A convenience sample of 29 subjects (10 men and 19 women) aged 39.4 ± 13.3 years completed two testing sessions on non-consecutive days (7.7 ± 3.2 days apart). Using the VALD ForceFrame we observed an excellent positive correlation for measurements of isometric trunk flexion strength (ICC 0.92; 95% CI 0.83-0.96) and isometric trunk extension strength (*ICC* 0.93; 95% CI 0.86-0.97) (Table 1). Measurement variations (SEM) were 13.99% and 13.72% for TF_ISO_ and TE_ISO_, respectively. Minimum detectable change results indicate that a change of 71.35N (7.28kg) represents a true change in isometric trunk flexion and a change of 101.20N (10.32kg) represents a true change in isometric trunk extension.

**Table 1.**
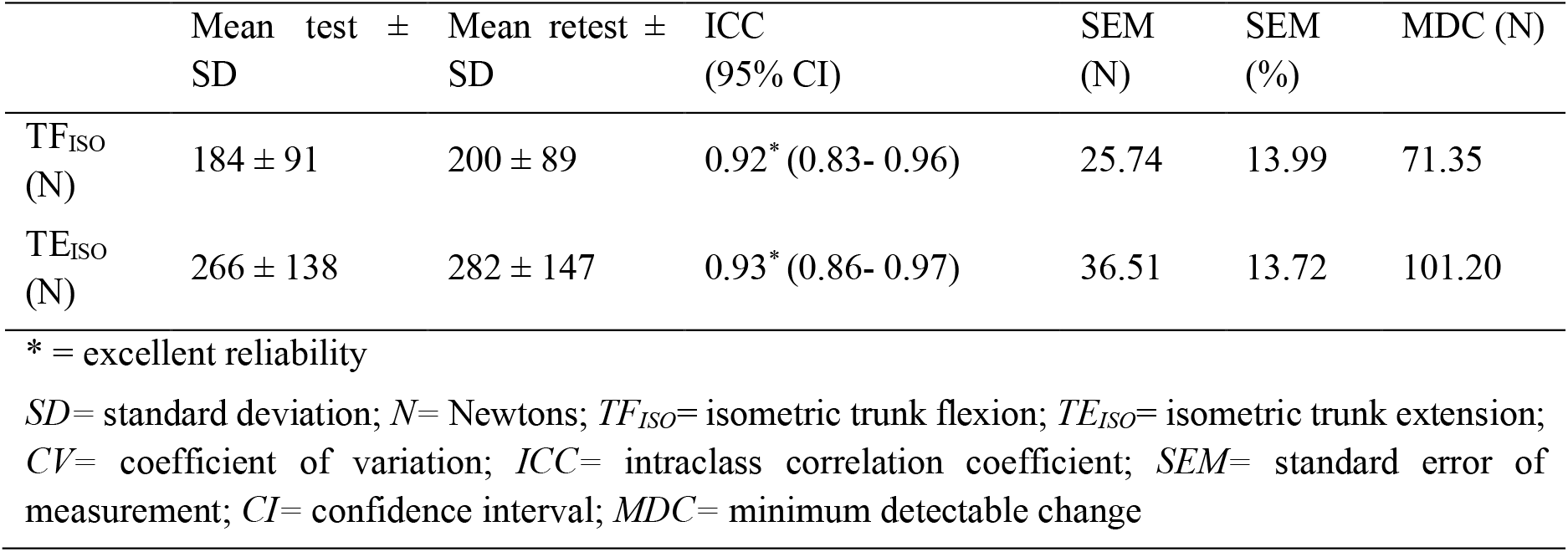
Reliability, Standard Error of Measurement and Minimum Detectable Change for Trunk Flexion and Trunk Extension Testing.

## DISCUSSION

Technological advances have led to the development of smaller, cheaper and ‘smarter’ sensors and interfaces, and these have found their way into many aspects of day to day life. There has been a rapid rise of integration of such devices into clinical measurement tools and this in turn has allowed clinicians to assess aspects of patient health and function in ways not previously possible. A reliable and accessible method of trunk strength measurement may afford clinicians an opportunity to more critically assess the effect of treatments aimed at improving trunk strength and control (such as an exercise programme aimed at improving multifidus function), and also to determine the impact of treatments that may interfere with lumbar muscle integrity (such as posterior lumbar surgery). Therefore, this study assessed the absolute and relative reliability of the VALD ForceFrame with the intention to measure trunk strength changes in individuals.

The results from this study indicate that the VALD ForceFrame demonstrates excellent test-retest reliability when used to measure isometric trunk flexion and extension strength (*r*= 0.92 and *r*= 0.93, respectively). These results are comparable to, or higher than, other studies that report on the reliability of fixed-frame devices for trunk strength testing (Guilhem et al., 2014; Müller et al., 2014; Harding et al., 2017; Roth et al., 2017; Cavuoto et al., 2019; Verbrugghe et al., 2019). One significant difference between the VALD ForceFrame and previously reported testing machines is the device infrastructure. Unlike the VALD ForceFrame, other devices are isokinetic in nature, which allow them to assess the rotational velocity and power of a muscle through a range of motion. Due to their cost and impracticality, such devices are scarcely used in out-patient clinical practice to deliver patients and practitioners with real-time feedback (Stark et al., 2011). These previously tested devices also require secure fixation at the hips, thighs and shoulder girdle so as to limit the reciprocal contraction of the synergist muscles. Previous studies have attributed improper patient stabilisation to poorer reliability outcomes (Moreland et al., 1997; Dvir and Prushansky, 2008). Contrary to this, the present study demonstrates that excellent test-retest reliability can be achieved by marking precise landmarks for force plate contact and through the secure fixation of the ankles alone. Moreover, the maximum MVC recorded in TE_ISO_ was 695N (70.87kg) and in TF_ISO_ 392N (39.97kg). Hand-held dynamometry would be unfeasible in this instance, as the results exceed 15kg and are deemed to require additional stabilisation (Brinkmann, 1994). Therefore, the authors conclude that the VALD ForceFrame is a hybrid testing system that reduces measurement error experienced with hand-held dynamometry and the impracticalities of isokinetic machines, to produce consistent findings of isometric trunk strength between testing sessions.

There are several different TF_ISO_ and TE_ISO_ testing positions described in the literature, the most of common of which are standing and upright sitting (Danneskiold-Samsøe et al., 2009; Guilhem et al., 2014; Müller et al., 2014; Harding et al., 2017; Roth et al., 2017; Cavuoto et al., 2019; Verbrugghe et al., 2019). An appropriate test position was chosen with consideration for the goals of the device; to minimise patient fixation in an effort to simplify testing procedures. Preliminary testing using electromyography revealed that, in standing and upright sitting positions, without fixation across the shoulder girdle and hips, there were large muscle contributions from the upper-back, shoulders, chest and hips. Therefore, a semi-flexed position (-40°) was adopted which minimised the contributions of other muscle groups and only required secure fixation at the ankles. The muscle activity of the trunk flexors at -40° were tested in a study by Deering et al. (2017) which revealed force production to be the greatest in this position, compared with trunk positions of 0°, -10°, -20° and -30°. This may explain why – when compared to healthy participants tested in upright sitting and standing– the TF_ISO_ measurements were largest in this study (Danneskiold-Samsøe et al., 2009; Guilhem et al., 2014; Roth et al., 2017).

The SEM and MDC values provide absolute measures of reliability (Weir, 2005). As we cannot control for all factors impacting the test scores, the SEM score quantifies the amount of error in the measurement. It provides a range as to where we would expect a score to fall, should we repeatedly perform the test. The SEM and SEM% for TF_ISO_ and TE_ISO_ was 25.74N (13.99%) and 36.51N (13.72%), respectively. Minimal detectable change (MDC) values indicate the smallest ‘real’ change that is not due to measurement error. The MDC of the trunk flexors was 71.35N and the MDC of the trunk extensors was 101.20N. That is, if there is a 71.35N change in trunk flexor Force or a 101.20N change in trunk extensor Force, these differences may be attributable to the effect of an intervention. The TF_ISO_ and TE_ISO_ SEM values in this study are up to 17.7Nm and 21.7Nm larger than in previous studies, respectively (Guilhem et al., 2014; Verbrugghe et al., 2019). This is expected as the SEM is calculated using the baseline standard deviation, of which is 89Nm and 147Nm in the current study. This is in comparison to standard deviation values reported in previous literature between 24.8Nm to 55.5Nm in TF_ISO,_ and, 60 to 89.3Nm in TE_ISO_ (Danneskiold-Samsøe et al., 2009; Guilhem et al., 2014; Müller et al., 2014; Deering et al., 2017; Harding et al., 2017; Verbrugghe et al., 2019), the upper limits of which are still significantly smaller to those reported here. While this suggests there is greater variation in our results, it is an anticipated finding provided the heterogenous population of participants in the present study. As the MDC is reliant on the SEM (MDC_95_= 1.96 x √2 x SEM), the MDC results in this study are understandably proportionally larger than those previously reported.

Despite the excellent test-retest reliability of the TF_ISO_ and TE_ISO_ tests, there are several limitations that warrant discussion. Firstly, while every effort was made by the examiner to standardise the trunk flexor position at -40°, without the use of secure fixation around the shoulder girdle it can be assumed that variations in the participants’ trunk angle occurred. Secondly, the larger trunk flexion averages reported in the current study have been attributed to the -40° testing position producing larger forces than upright positions (Deering et al., 2017). However, recording participants’ weight would have detailed more about the population, and may have explained why the forces produced in this study were larger than in others. Lastly, participants represented a convenience sample of family members, friends and colleagues. Caution should be taken when generalising the results of this study to other populations.

## CONCLUSIONS

This study shows that a fixed-frame dynamometer demonstrates excellent test-retest reliability when used for testing isometric trunk flexion and extension strength in community-dwelling adults. It justifies the integration of a convenient and portable dynamometry system with limited secure fixation into clinical practice, that overcomes the clinician error often reported with hand-held dynamometry. Using the VALD ForceFrame muscle strength changes may be efficiently and accurately detected, providing real-time feedback for patients and practitioners on musculoskeletal status.

## Data Availability

The datasets generated and analysed during the current study are not publicly available due to their potentially identifiable nature and participant confidentiality requirements. De-identified data may be available from the corresponding author on reasonable request, subject to ethical approval and applicable institutional requirements.

## ACKNOWLEDGEMENTS

The authors would like to thank the staff at the Brisbane Private Hospital, friends and family who volunteered their time to be tested for this study. The authors would like to declare that there are no conflicts of interest to disclose, and that this research was supported by a research grant from Johnson & Johnson Health Care Products & Pharmaceuticals (grant number: DPS-SPINE-2020-017).

